# Sparse Parallel Independent Component Analysis and Its Application to Identify Stable and Replicable Imaging-genomic Association Patterns in UK Biobank

**DOI:** 10.1101/2022.06.27.22276981

**Authors:** Kuaikuai Duan, Jiayu Chen, Zening Fu, Rogers F. Silva, Vince D. Calhoun, Michela Dell’Orco, Nora I Perrone-Bizzozero, Yuhui Du, Wenhao Jiang, Jingyu Liu

## Abstract

Data fusion analyses of brain imaging and genomics enable the linking of genomic factors to brain patterns. Due to the small to modest effect sizes of common genetic variants, it is usually challenging to reliably identify relevant genetic factors from the rest of the genome with the typical sample size in neuroimaging studies. To alleviate this problem, we propose sparse parallel independent component analysis (spICA) to leverage the sparsity of individual genomic sources, building upon the existing parallel independent component analysis (pICA) algorithm. Sparsity is enforced by performing Hoyer projection on the estimated independent sources. Simulation results demonstrate that spICA yields improved recovery of imaging-genomic associations and sources compared to pICA. We applied spICA to whole-brain gray matter volume (GMV) and whole-genome single nucleotide polymorphisms (SNPs) data of five different sets of 24,985 discovery samples in the UK Biobank. We identified three GMV sources significantly and stably associated with one SNP source and replicated these associations. GMV sources highlighted frontal, parietal, and temporal regions. Their corresponding loadings on individuals were related to multiple cognitive measures, and the temporal region interacts with age influencing cognition. The SNP component underscored SNPs in chromosome 17 that were enriched in the inflammation response pathway and in regulation effect in the prefrontal cortex via gene expression, methylation, transcription expression, and isoform percentage.

## I. INTRODUCTION

ADVANCEMENTS in brain imaging and multi-omics techniques promote the field of imaging genomics that aims at delineating genomic underpinnings of neurobiological alterations related to brain disorders, which will help better understand the pathology of diseases and potentially aid (personalized) early intervention planning. However, achieving this goal has proven challenging given the complexity of disorders, the large dimensionality (millions) of imaging and genomics features in comparison to practical sample sizes (hundreds), and modest effect sizes of individual genetic variants revealed by genome-wide association studies (GWAS) [1-4]. Data-driven multivariate fusion and sparse analyses can help tackle these challenges, since multivariate fusion methods [5] aggregate related variables with small effect sizes into one network/source and simultaneously identify brain imaging-genomic associations at the network level to boost statistical power, and sparsity optimization suppresses the background/noise and enhances the signal of interest.

Several data-driven multivariate (sparse) fusion approaches have been proposed for imaging genomics, including sparse canonical correlation analysis (sCCA) [6, 7], sparse partial least squares (sPLS) [8], sparse reduced rank regression (sRRR) [9], and parallel independent component analysis (pICA) [10, 11]. sCCA, sPLS, and sRRR are extensions of CCA, PLS, and RRR, respectively, which implement a sparsity constraint exploiting *l*_1_ norm to alleviate uncontrolled overfitting in the original methods to some extent. CCA and PLS maximize the correlation and covariance between two modalities, respectively. RRR minimizes the error of the multivariate regression by reducing the rank of the projection matrix mapping the input to the output. We note pICA differs from the aforementioned approaches in that it prioritizes extracting maximally independent components from individual modalities, while simultaneously optimizing the correlations between multimodal component expressions. The advantages and limitations of each method have been reviewed elsewhere [5]. sCCA, sPLS, and sRRR are limited to decomposing the same number of factors/components for all modalities, while pICA allows different numbers of components for different modalities. Moreover, when the intrinsic dimensionality of a modality is much larger than the sample size, especially with genomic single nucleotide polymorphism (SNP) data, the search space becomes so large that even methods such as sCCA, sPLS, and sRRR are susceptible to overfitting. The very large dimensionality condition also makes it difficult for pICA to separate signals of interest (e.g., SNPs involved in a specific brain alteration) from the background genomic profile. Since the variance of genotypes carried by SNPs of interest is typically comparable to, or slightly higher than, that of background SNPs (i.e., SNPs modulating other or unknown biological processes), the independent components from pICA tend to assign similar or slightly higher weights for SNPs of interest compared to background SNPs [12]. In this study, we propose a sparsity regulated pICA to mitigate issues with signal-background-separation. Like pICA, the use of independence leverages higher-order statistics to overcome overfitting issues, while (nonlinear) sparsity regularization yields a crucial “denoising” effect for better signal detection.

Among sixteen sparsity measures that have been summarized in [13], we selected the Hoyer index as the most promising, since it satisfies five out of six desirable axiomatic attributes of sparsity measures while also being differentiable and, thus, amenable to optimization. The Hoyer index is an extension of the ratio between *l*_1_ and *l*_2_ norms and can achieve any desired level of sparseness. Comparing to *l*_1_ norm, Hoyer index is scale invariant, which is well suited for ICA frameworks since the scale of decomposed components is arbitrary. Our previous work [12] has demonstrated that incorporating nonlinear Hoyer projection directly on independent SNP sources estimated from infomax ICA improved the SNP component detection accuracy under various scenarios. Moreover, enforcing Hoyer projection directly while maximizing independence of the sources enables explicit (rather than implicit) adjustment of sparseness of sources, which saves trial-and-error for finding parameters yielding the desired degree of sparseness. Thus, we propose sparse parallel ICA (spICA) to incorporate nonlinear sparsity optimization directly on sources estimated from the pICA framework, not only taking advantage of correlation optimization but also leveraging the desirable sparsity properties of the Hoyer index for nonlinear denoising. The effectiveness and the robustness of spICA are examined with simulated imaging and genetic data.

To examine the capability of the proposed spICA in real data, we used whole-brain gray matter volume (GMV) and whole-genome SNP array data collected from 35692 European Caucasians recruited in the UK Biobank (UKB, http://www.ukbiobank.ac.uk) project. To assess the stability of spICA-identified GMV-SNP component pairs, we applied spICA to five randomly generated discovery datasets (see details later) separately to identify significantly correlated GMV-SNP component pairs and evaluate the similarities of identified spatial maps among five spICA results. The replicability of spICA-identified GMV-SNP component pairs is examined using each corresponding replication dataset. Specifically, we randomly partitioned full samples five times to generate five discovery and replication datasets, where each discovery and replication datasets contained 24985 (70% full samples) and 10707 participants (the remaining 30% samples), respectively. The imaging-genetic pairs are considered stable and replicable if they are consistently significant and with the same inferences in more than three discovery datasets (i.e., ≥ 60%) and replicated in the corresponding replication datasets. Moreover, we also investigated the biological relevance of the GMV and SNP components as well as their relationship to cognitive and neuroticism.

## II. METHODS AND MATERIALS

The spICA proposed here is an extension of pICA [10], which imposes additional sparsity constraints on components/sources. The goal of pICA is to maximize component independence for each modality in parallel (i.e., multiple ICAs) while simultaneously enhancing the intercorrelation between specific loadings across modalities. In spICA, the sparsity constraint is enforced at each iteration by applying the Hoyer projection [12, 14] on the estimated components directly, which enhances signals of interest and effectively suppresses background information. Explicit adjustment of sparseness of sources avoids trial-and-error for finding parameter settings in gradient optimization to achieve desired sparseness of sources.

### A. Sparse Parallel Independent Component Analysis

The cost function of the proposed spICA is shown in (1). spICA maximizes the sum of differential entropies (H(**y**_1_) + H(**y**_2_)) and square of the intercorrelation between modalities (corr^2^(**a**_1_, **a**_2_)) with respect to unmixing matrices (**W**_1_, **W**_2_), subject to Hoyer index constraints on the components of both modalities (Hoyer(**s**_1_) ≥ q_1_, Hoyer(**s**_2_) ≥ q_2_).

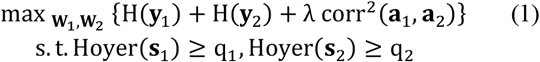

where

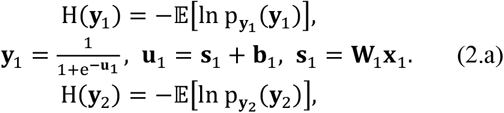

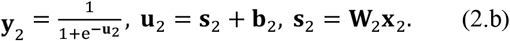

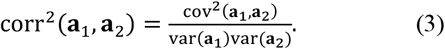

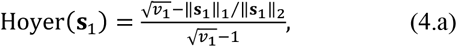

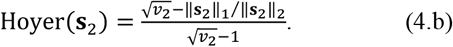

where **X**_1_, **X**_2_ are input data from two modalities with the dimension of subject-by-variable (voxels or SNP loci). Data are modeled as linear mixtures of independent components, i.e., **X**_1_ **= A**_1_**S**_1_, **X**_2_ **= A**_2_**S**_2_. Mixing matrices, **A**_1_ and **A**_2_, have subject-by-component dimensionality and can be estimated as 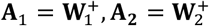 respectively, where ^+^ is the pseudoinverse operation. Independent components, **S**_1_and **S**_2_, are component-by-variable (voxels or SNP Loci), and can be estimated as **S**_1_ **= W**_1_**X**_1_ and **S**_2_ **= W**_2_**X**_2_, respectively. Each row of **S**_1_ or **S**_2_ is one independent component (**s**_1_, **s**_2_, respectively), with component weights reflecting the relative involvement of variables (voxels or SNP loci) in the component. Each column of **A**_1_or **A**_2_ is a loading vector (**a**_1_, **a**_2_, respectively) and their values represent expression levels of the corresponding component across subjects. p_**y**1_ (**y**_1_), p_**y**2_ (**y**_2_) are probability density functions of output vectors **y**_1_, **y**_2_, computed as nonlinear transformations of bias-adjusted sources **u**_1_, **u**_2_, respectively. corr(⋅), cov(⋅), and var(⋅) are correlation, covariance, and variance operators, respectively. *v*_1_ and *v*_2_ are the numbers of variables (voxels or SNP loci) in components **s**_1_, **s**_2_, respectively. ‖⋅‖_1_ and ‖⋅‖_2_ are the *l*_1_ and *l*_2_ norm operators, respectively. λ is a regularizer to balance between independence and correlation maximization. λ is initialized as one and is adaptively reduced by a factor of 0.9 whenever the slope of the entropy of components (H(**y**_**1**_) and H(**y**_**2**_)) smaller than −10^−5^ is detected. *q*_1_, *q*_2_ are threshold parameters for the Hoyer sparsity index, and 𝔼[⋅] is the expectation operator. The value of Hoyer index is between 0 and 1. The larger the Hoyer index, the sparser the source.

Maximizing the independence of components **s**_1_, **s**_2_ is achieved by maximizing the entropy of output vectors **y**_1_, **y**_2_ using infomax [15]. This is solved by stochastic gradient descent using the relative/natural gradient 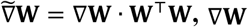 is the traditional gradient) [16] of H(**y**_1_) and H(**y**_2_) with respect to **W**_1_ and **W**_2_, and the traditional gradient of H(**y**_1_) and H(**y**_2_) with respect to **b**_1_ and **b**_2_, respectively [17]:

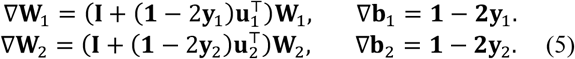

where **I** is an identity matrix, and **1** is a column vector of ones. ⊤ is the matrix/vector transpose operator. Correlation optimization is solved by gradient descent using the traditional gradient of corr^2^(**a**_1_, **a**_2_) with respect to **A**_1_ and **A**_2_, respectively [10]:

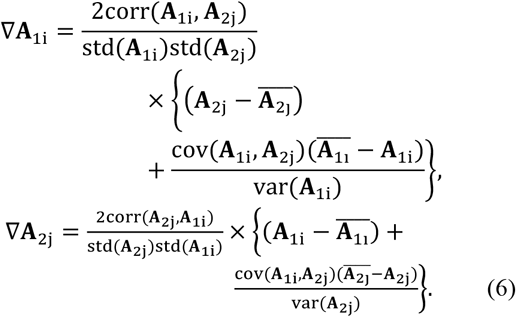

where **A**_1i_ and **A**_2j_ represent the loading coefficients of the i-th component of modality one and the j-th component of modality two, respectively. std(.) denotes the standard deviation, ⨪ represents an average operation. We set the number of pairs to be optimized as one in this study to avoid overestimating the correlation. So, only the loading coefficient pair with the strongest correlation is optimized for correlation enhancement at each iteration. Sparsity constraints are solved by applying the Hoyer projection to the estimated components **s**_1_ **= w**_1_**X**_1_, **s**_2_ **= w**_2_**X**_2_ when their Hoyer indices are less than the preset thresholds *q*_1_, *q*_2_. The Hoyer projection is a nonlinear transformation and is implemented according to the equations above and as described in Hoyer’s paper [14]. During Hoyer projection, the *l*_2_ norm of the components are unchanged, while the *l*_1_ norm are updated to achieve the desired Hoyer indices *q*_1_ and *q*_2_ according to (4.a) and (4.b). The desired Hoyer index value depends on the data properties of each modality, such as number of variables (voxels or SNP loci), the extent of signal regions, as well as expected minimum signal-to-background ratio (SBR) as defined in [12] (the larger the SBR, the easier to separate signal from background). The preset thresholds *q*_1_, *q*_2_ can be estimated from simulated components with an expected (*a priori*) component SBR for each modality, assuming that the background and signal regions of each component form logistic (mean = 0, variance =3) and Laplacian (mean = expected SBR, scale factor = 1) distributions, respectively. Additionally, we conservatively initialize the step sizes for sparsity enhancement to 5 ⋅ 10^−3^ (for sMRI) and 5 ⋅ 10^−4^ (for SNP) and, in order to prioritize independence maximization, dynamically reduce them by a factor of 0.98 during optimization whenever the slope of the entropy smaller than −10^−5^ is detected.

Fig. 1 shows the flow chart of the proposed spICA, where the dashed boxes and dashed lines indicate extra steps with respect to pICA, and red feedback arrows indicate updates which occur last in each iteration. At the beginning, we initialize the input data **X**_i_, **X**_g_, threshold parameters for the Hoyer sparsity index q_i_, q_g_, learning rates for independence *l*_i_, *l*_g_, step sizes for sparsity enhancement Δ_hi_, Δ_hg_ for imaging and genetic modalities, separately. Secondly, the unmixing matrices **W**_i_, **W**_g_ are updated based on infomax using the relative gradient of differential entropies **H**(**s**_i_), **H**(**s**_g_) with respect to the unmixing matrices **W**_i_, **W**_g_, respectively. If the Frobenius norm of the difference between the current and previous unmixing matrices is less than a threshold, then the algorithm converges, and the optimization stops for this modality. Otherwise, successively update the component and unmixing matrices using Hoyer projection and correlation optimization, respectively. It is worth noting that pICA optimizes the independence and correlation of components through **W**_i_, **W**_g_ updates, while sparsity optimization is directly on the components. To take full advantage of sparsity optimization and successfully propagate its effect, we update the data **X**_i_ and **X**_g_ for the next iteration by reconstructing it from the sparsity-optimized component and the current unmixing matrix from infomax. The pseudocode of the proposed spICA is provided in the supplemental materials.

**Fig. 1.**
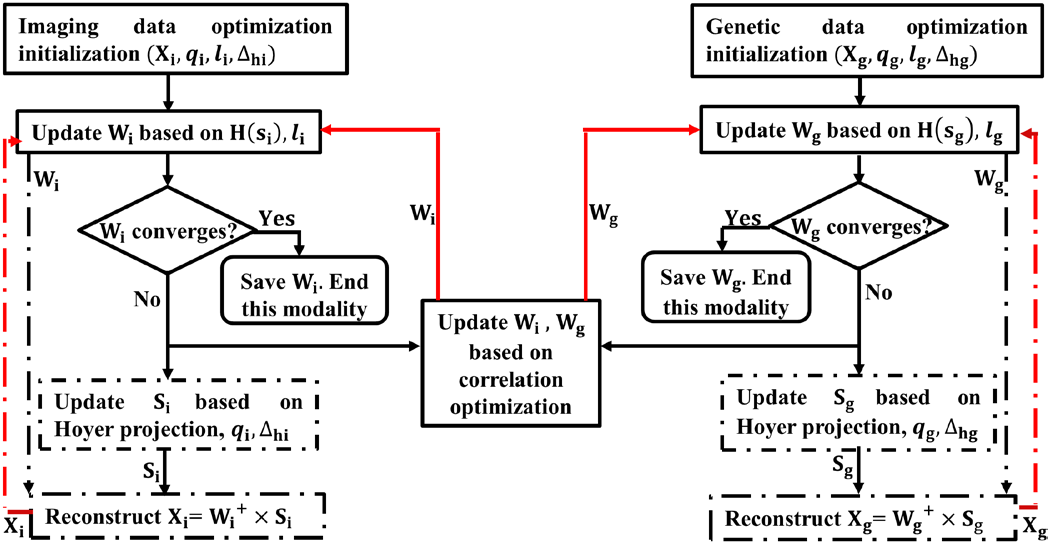
The flow chart of spICA. Note, the dashed boxes and dashed lines indicate extra steps with respect to pICA (red feedback arrows indicate updates which occur last in each iteration). Sparsity is optimized on each modality separately through updating source ***S***_***i***_ or ***S***_***g***_ based on Hoyer projection, which is further propagated to the next iteration by reconstructing the data ***X***_*i*_ or ***X***_*g*_ with sparsity-enhanced source.

### B. Simulation

We simulated structural MRI (sMRI) and SNP data to compare the proposed spICA and its sparsity-free alternative (i.e., pICA) for their capability to extract accurate and sparse sMRI and SNP sources/components, and recover the designed linkages/associations between sMRI and SNP component under four scenarios: (1) Varying sMRI-SNP correlation with other variables fixed; (2) Varying number of subjects with other variables fixed; (3)Varying number of SNPs of interest (i.e., the extent of signal region) with other variables fixed; (4) Varying SNP pattern strength and white Gaussian noise superimposed on sMRI components with other variables fixed. Details of four scenarios were described as follows:

#### 1) Scenario 1

Varying sMRI-SNP correlation, setting number of subjects = 200, SNP pattern strength (defined below) = 3.5, number of SNPs of interest = 150 (total number of SNPs = 10000), peak signal-to-noise ratio (PSNR) of white Gaussian noise = 21dB (for sMRI), sMRI sparsity parameter *q*_*i*_ = 0.85 and SNP sparsity parameter *q*_*g*_ = 0.4 (to match ground-truth Hoyer index mentioned in sMRI and SNP data simulation sections, the same for scenarios 2-4). Correlation between sMRI and SNP varied from 0.15 to 0.4 with a step size of 0.05. SNP pattern strength is defined as:

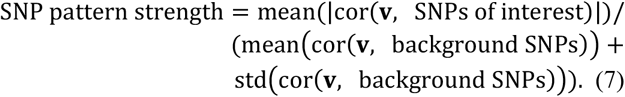

Here, **v** is the representative SNP that best resembles the case-control status and is selected from the set of SNPs of interest (this representative SNP is excluded from the SNPs of interest when estimating SNP pattern strength). cor(.) and std(.) denote correlation and standard deviation operations, respectively.

#### 2) Scenario 2

Varying number of subjects, setting sMRI-SNP correlation = 0.25, SNP pattern strength = 3.5, number of SNPs of interest = 150 (total number of SNP = 10000), PSNR of Gaussian noise = 21dB, sMRI sparsity parameter *q*_*i*_ = 0.85 and SNP sparsity parameter *q*_*g*_ = 0.4. Number of subjects varied from 100 to 1000 (i.e., 100, 200, 300, 500, 700, 1000).

#### 3) Scenario 3

Varying number of SNPs of interest, setting number of subjects = 200, sMRI-SNP correlation = 0.25, SNP pattern strength = 3.5, total number of SNPs = 10000, PSNR of Gaussian noise = 21dB. sMRI sparsity parameter *q*_*i*_ = 0.85. Number of SNPs of interest varied from 50 to 500 (i.e., 50, 100, 150, 500). Hoyer indices of these four SNPs of interest settings were estimated around 0.4. Thus, we set SNP sparsity parameter *q*_*g*_ = 0.4.

#### 4) Scenario 4

Varying SNP pattern strength and levels of white Gaussian noise on sMRI component matrix, setting number of subjects = 200, sMRI-SNP correlation = 0.25, number of SNPs of interest = 150 (total number of SNP = 10000), sMRI sparsity parameter *q*_*i*_ = 0.85 and SNP sparsity parameter *q*_*g*_ = 0.4. White Gaussian noise with PSNR varying from 7 dB to 25 dB were superimposed on the sMRI components, and different strengths (2, 4, and 5) were employed in the SNP patterns. All combinations of PSNR and SNP pattern strength were evaluated.

Note that for each scenario, 10 sets of white Gaussian noise were generated. Thus, spICA and pICA were run on 10 sets of sMRI data. The performance measures were the average across ten runs, and the corresponding standard errors were estimated across ten runs.

#### 5) sMRI data simulation

sMRI data ***X***_*i*_ (dimension: subject-by-voxel; number of voxels: 31064; under scenarios 1, 3, and 4, number of subjects = 200; for scenario 2, number of subject varied from 100 to 1000) were simulated [12] as the product of a loading matrix ***A***_*i*_ (dimension: subject-by-component, number of components: 7) and a component matrix ***S***_*i*_ (dimension: 7 components by 31064 voxels) containing seven non-overlapping brain regions (Fig. 2 (a)) generated by the simTB toolbox [18]. All components were designed to have a ground-truth Hoyer index around 0.85. Without loss of generality, the loadings of the first sMRI component were designed to be correlated with the loadings of the first SNP component. The loadings of the remaining six sMRI components were sampled from a uniform distribution 𝒰(0,1), yielding sample correlations much lower than the designed correlation between modalities. For scenarios 1-3, white Gaussian noise with PSNR of 21dB was superimposed on the component matrix to yield noisy components. For scenario 4, white Gaussian noise with PSNR ranging from 7dB to 25dB at a step size of 2dB was superimposed on the component matrix to yield noisy components. The smaller the PSNR, the heavier the noise, thus the more difficult to recover the sources.

**Fig. 2.**
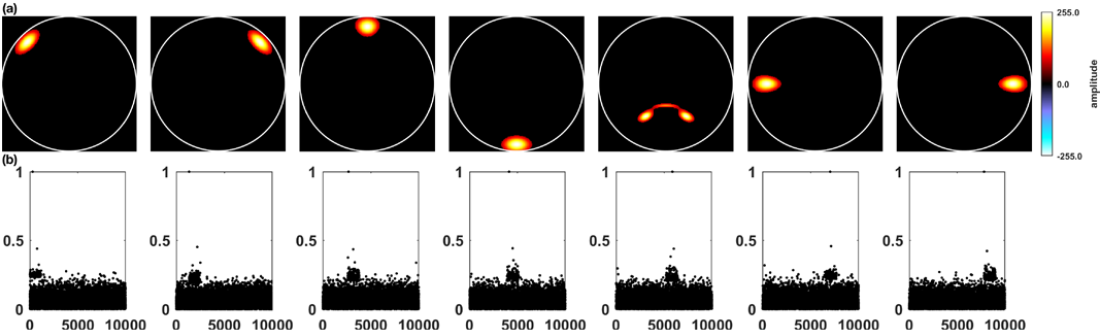
(a) Seven ground-truth sMRI components, (b) Correlation between the representative SNP and the seven nonoverlapping SNP component patterns (SNP pattern strength = 3.5).

#### 6) SNP data simulation

SNP data ***X***_*g*_ (dimension: subject-by-SNP; number of SNPs: 10000; under scenarios 1, 3, and 4, number of subjects = 200; for scenario 2, number of subject varied from 100 to 1000) were generated using the simulate function in PLINK (http://zzz.bwh.harvard.edu/plink/), which were designed to contain 7 nonoverlapping sparse genetic patterns (Fig. 2(b)) with randomly generated case-control status matrix. Each pattern was a realization of one genetic component in the population, where SNPs with higher correlations with the representative SNP (the SNP closely resembles case-control status) were the SNPs of interest and those with lower correlations with the representative SNP were background SNPs. All simulated SNPs were unrelated and in linkage equilibrium. Under scenarios 1, 2, and 4, 150 loci were set as the SNPs of interest for each genetic pattern to resemble the sparsity of genetic data. Under the scenario 3, SNPs of interest for each genetic pattern varied from 50 to 500 (50, 100, 150, and 500). For all four scenarios, the Hoyer indices of such simulated SNP patterns were estimated around 0.4. The case-control status matrix was treated as a proxy of the loading matrix for the SNP data. Under each scenario, the strength of seven SNP patterns were the same.

### C. spICA Application to UKB Imaging Genomics

The UKB is a population-based adult study that collected deep genetic and phenotypic information from ∼500,000 individuals, and neuroimaging data from ∼100,000 individuals recruited across the United Kingdom [19, 20]. The study was approved by the Research Ethics Committee of UKB. Data in the current study were obtained under UKB application 34175. sMRI and SNP data from 35,975 European Caucasians were included in this study.

#### 1) sMRI data preprocessing

T1-weighted MRI images were collected at three imaging centers (Cheadle, Reading, and Newcastle) with identical scanners (3T Simens Skyra), which were free from major software or hardware updates throughout the study [21]. In brief, the scanning parameters were TI/TR = 880/2000 ms, resolution = 1×1×1mm, sagittal acquisition, FOV = 208×256×256. The included T1-weighted images were segmented into six types of tissues (gray matter, white matter, cerebrospinal fluid, bone, soft tissue, and air/background) using SPM 12 with the default tissue probability map (TPM). Subsequently, gray matter images were normalized into Montreal Neurological Institute (MNI) space using the default spatial normalization function in SPM 12, with resolution = 1.5×1.5×1.5mm. Normalized gray matter images were then modulated with the Jacobian determinants of the nonlinear transformation and smoothed with a 6×6×6mm^3^ Gaussian kernel. Further quality control was conducted to retain samples with a correlation larger than 0.9 with the mean gray matter map across all the individuals, yielding 35,692 subjects. GMV refinement (i.e., masking) selected voxels with mean GMV larger than 0.2, yielding 416742 voxels for further analyses.

#### 2) SNP data preprocessing

SNP data were genotyped using the Applied Biosystems UK BiLEVE Axiom Array by Affymetrix (for 3465 samples) or a closely related Applied Biosystems UK Biobank Axiom Array (for 32227 samples), where 95% markers were shared between these two arrays [22]. Genotyped SNPs were phased and imputed to ∼96 million variants using the Haplotype Reference Consortium resource and the merged UK10K and 1000 Genomes phase 3 reference panels [22]. Samples with mismatches between self-reported sex and genetically derived sex were excluded. We further restricted our analyses to unrelated European Caucasians. SNP variants with imputation r^2^ ≤ 0.3, missingness rate ≥ 5%, minor allele frequency ≤ 0.01, Hardy–Weinberg Equilibrium p value ≤ 1×10^−6^ were filtered out. Subsequently, linkage-disequilibrium based pruning with r^2^ = 0.5 and window size = 50kb was applied to filter out highly dependent variants, resulting in 1,151,605 SNPs for further analysis. The first 20 principal components of SNP data, potentially accounting for population stratifications [23-25] that are not our research interest, were excluded before spICA. After QC for both GMV and SNP data, 35,692 uncorrelated European Caucasians (18835 females, 63.69±7.51 years old) were retained for further analyses. Cheadle, Reading, and Newcastle centers scanned 22151, 4528, and 9013 participants, respectively.

#### 3) Applying spICA to preprocessed sMRI and SNP Data and evaluating stability and replicability

The proposed spICA was applied to GMV and SNP data of each of five sets of discovery samples produced by randomly partitioning the full set of samples five times (in each partition, 70% samples (24985 participants) for discovery and the remaining 30% for replication). For each spICA trial, the component number of GMV data was set as 25 to be consistent with Rodrigue’s ICA-based UKB imaging-genetic study [24]. SNP component number was set as 50 to retain large-scale SNP networks. The regularizer λ was initialized as one and the Hoyer constraint threshold of SNP data was estimated as 0.45 (i.e., *q*_*g*_ = 0.45). No sparsity constraint was imposed on GMV data (since imaging sources are usually easy to separate, we set *q*_*i*_ = 0 so that spICA would not attempt to optimize the sparsity for GMV data). GMV-SNP associations were evaluated with the regression model (a): loadings of a GMV component = loadings of a SNP component + age + gender + site + genotype array, where all predictors were treated as fixed effect. Bonferroni at p < 0.05 was applied to correct for 1250 (25×50) GMV-SNP association tests for each spICA result. The generalizability of identified GMV-SNP pairs from each spICA result was validated with the corresponding replication dataset (10707 participants). Loadings of highlighted GMV and SNP components in replication dataset were computed with a projection method (supplemental S1 for details of projecting exact same components into new data) [26], and corresponding GMV-SNP associations were evaluated with the abovementioned model (a). GMV-SNP pairs are considered as replicated if the association p values for the replication dataset pass false discovery rate at p < 0.05 correction for pairs that survived the correction test in the discovery dataset. Spatial map similarities between the identified GMV and SNP components from each of the five discovery datasets were assessed using Pearson correlation tests.

Finally, to assess the stability of the results, we need to determine the base partition first. The base (or “final”) partition was selected from one of the five partitions based on their GMV-SNP associations and spatial map similarities (spatial maps with correlation |*r*| > 0.75 were considered as similar) across the five discovery datasets using two strategies: (1) if ≥ 3 partitions (i.e., ≥ 60%) yielded similar GMV-SNP pairs, we randomly selected one partition from those generating similar GMV-SNP pairs as the base (or “final”) partition; (2) if less than 3 partitions yielded similar GMV-SNP pairs, we considered the results being unstable, and no further stability and replicability needed. After determining the base (or “final”) partition, we then evaluate the stability of GMV-SNP pairs by examining (1) whether these pairs were significantly linked in the discovery datasets and replicated in the replication datasets for the remaining four partitions; (2) how do the linked GMV and SNP components in the remaining four partitions similar to those in the base partition.

#### 4) Cognitive and neuroticism relevance of the identified GMV and SNP components

To better understand the identified GMV and SNP components, we investigated their involvements in cognition and neuroticism. The cognitive domains assessed in UKB included numeric memory, verbal-numerical reasoning, reaction time, and visual memory [27]. The cognitive variables included in this study as suggested by Lyall et. al.[27] were maximum digits remembered correctly (numeric memory), fluid intelligence (verbal-numerical reasoning), mean time to correctly identify matches (reaction time), number of incorrect matches in round (visual memory). Neuroticism, a depression-related personality trait, was evaluated based on 12 neurotic behavior domains from the Eysenck Personality questionnaire [28]. Cognitive performance measured at the time of sMRI scan were used in this study (see Supplemental Table S1 for details). Associations between identified GMV components and cognitive measurements/neuroticism were tested using the model: a cognitive measure/neuroticism = loadings of a GMV component + age + gender + site. SNP component association with cognitive measurements/neuroticism was evaluated with the model: a cognitive measure/neuroticism = loadings of a SNP component + age + gender + genotype array. Multiple comparisons were corrected at FDR at p < 0.05.

#### 5) Highlighted brain regions, SNP loci and their biological relevance

Each of the identified GMV and SNP components were first normalized to have a zero mean and unit standard deviation. Highlighted regions of each identified GMV component were selected with |*z*| > 2.5 and mapped into the Talairach atlas [29]. Top SNPs of the identified SNP component were selected with |*z*| > 3.5. The biological relevance of top SNPs was further examined including (i) gene annotation and enrichment in biological functions, pathways and diseases using Ingenuity Pathway Analysis (IPA, QIAGEN Inc., https://www.qiagenbioinformatics.com/products/ingenuity-pathway-analysis) and Gene Ontology (http://geneontology.org/), (ii) regulation effects on gene/transcript expression, isoform percentage, and DNA methylation in the prefrontal region based on the summary statistics of eQTL, tQTL, and isoQTL available on the PsychENCODE website (http://resource.psychencode.org/) and mQTL based on Jaffe’s study [30]. Enrichment of eQTL, tQTL, isoQTL and mQTL were evaluated by random selecting the same number (determined by top SNPs highlighted by spICA) of SNPs from the whom genome 10000 times and assessing how many selections yielded to numbers of eQTL, tQTL, isoQTL and mQTL larger than that of spICA-identified top SNPs.

## III. Simulation Results

The proposed spICA and its sparsity-free alternative, pICA, were tested and evaluated using carefully designed simulated sMRI and SNP data under four scenarios that most variables were fixed and only one variable varied under each scenario (see details in Method section B). Performance measures include the recovered correlation value of the designed sMRI-SNP pair, correlations between recovered sMRI components and ground-truth, correlations between recovered SNP components and designed SNP component patterns, and Hoyer indices (measuring sparsity) of recovered sMRI and SNP components.

Fig. 3 displays the recovered sMRI-SNP correlations under scenario 1 with the ground-truth correlations ranging from 0.15 to 0.4. Both pICA and spICA largely recovered the designed sMRI-SNP correlations. Varying sMRI-SNP correlations had negligible effects on the accuracy and sparsity of recovered sMRI and SNP components. Thus, they are averaged across different correlation settings. The average correlation between recovered sMRI components and ground-truth ones from pICA and spICA were 0.73 and 0.96, respectively. Recovered SNP components had average correlation with ground-truth ones of 0.42 and 0.46 for pICA and spICA, respectively. The Hoyer index of recovered sMRI components from pICA and spICA were 0.36 and 0.79, respectively. Recovered SNP components had Hoyer indices of 0.24 and 0.27 for pICA and spICA, respectively. Altogether, spICA yielded more accurate and sparser sMRI and SNP components compared to pICA. The improvement for sMRI components is more obvious compared to SNP components, where the improvement for SNP components may be limited by the designed challenging case (i.e., SNP pattern strength (see the definition in Method) = 3.5).

**Fig. 3.**
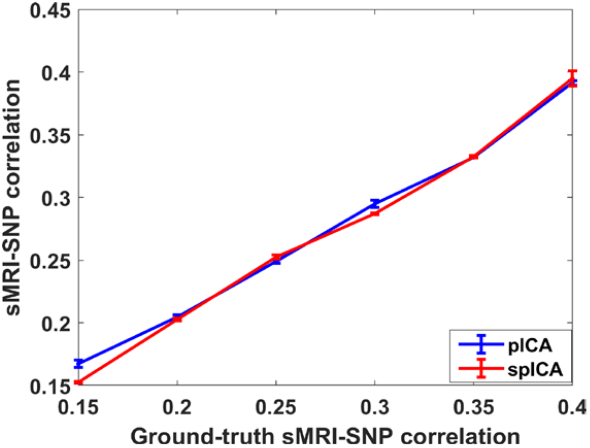
Scenario 1: The sMRI-SNP correlations detected by spICA (red) and pICA (blue) while varying ground-truth sMRI-SNP correlations from 0.15 to 0.4. Note, color representations are consistent for all simulation results (Fig. 3-7). Overall, both pICA and spICA recovered designed sMRI-SNP correlations, and spICA recovered sMRI components with much higher accuracy and sparsity (not shown), and consistently higher SNP component accuracy and sparsity compared to pICA.

Fig. 4 plots the recovered sMRI-SNP correlations and the accuracy of recovered SNP components under scenario 2 with number of subjects varying from 100 to 1000 and sMRI-SNP correlation fixed at 0.25 (closely matching real-world scenarios). Fig. 4(a) shows that pICA and spICA recovered the designed sMRI-SNP correlation when number of subjects was larger than 100. When the number of subjects was 100, both spICA and pICA failed to recover the designed sMRI-SNP correlation of 0.25. Fig. 4(b) reflects that the accuracy of recovered SNP components increased as number of subjects increased, and spICA always yielded higher accuracy compared to pICA. Varying number of subjects had negligible effect on recovery of sMRI components and sparsity of sMRI and SNP components (not plotted). The average correlation between recovered sMRI components and ground-truth ones across different settings for pICA and spICA were 0.73 and 0.95, respectively. The Hoyer indices of recovered sMRI components from pICA and spICA were 0.36 and 0.78, respectively. Recovered SNP components had Hoyer indices of 0.25 and 0.28 for pICA and spICA, respectively.

**Fig. 4.**
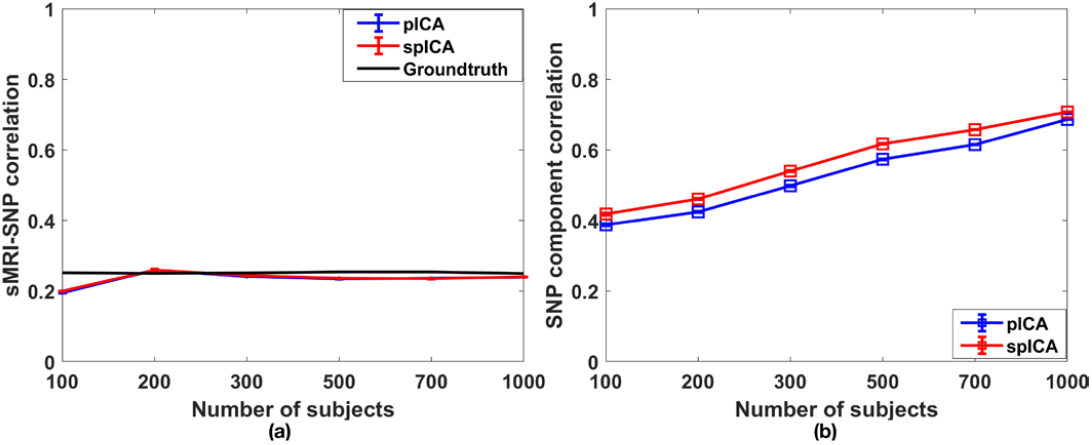
Scenario 2: Varying number of subjects from 100 to 1000, the recovered (a) sMRI-SNP correlations (the ground-truth was 0.25 (black)) and (b) SNP component accuracy by spICA (red) and pICA (blue). Note, standard errors of SNP component correlation were close to 0 and are difficult to visualize in the figure (same for Fig 5 and Fig 7). Overall, spICA recovered sMRI components with much higher accuracy and sparsity (not shown), and consistently higher SNP component accuracy and sparsity compared to pICA. For SNP components, pICA required 100 subjects more to attain similar accuracy of spICA.

Fig. 5 displays the recovered sMRI-SNP correlation and accuracy and Hoyer value of recovered SNP components under scenario 3 with number of SNPs of interest varying from 50 to 500 and sMRI-SNP correlation fixed at 0.25. Fig. 5(a) shows that pICA and spICA recovered the designed sMRI-SNP correlation when number of SNPs of interest was larger than 50. Fig. 5(b) indicates that the recovery accuracy of SNP components increased as number of SNPs of interest increased, and spICA yielded higher accuracy compared to pICA when number of SNPs of interest was larger than 50. Fig. 5(c) demonstrated that spICA improved the sparsity of SNP components compared to pICA. Fig. 5(b) and (c) showed that with 200 subjects and SNP pattern strength of 3.5, both pICA and spICA failed to recover the designed sMRI-SNP correlation and SNP component patterns, when the number of SNPs of interest was 50. This result indicates (s)pICA methods require a minimum ratio between the number of SNPs of interest and the total number of SNPs analyzed. The trend that the sparsity increased as the number of SNPs of interest increased is the result of increased accuracy of SNP components (the more accurate the recovered SNP component, the closer the recovered Hoyer index is to the ground-truth). Varying number of SNPs of interest had negligible effect on recovery and sparsity of sMRI components. The average correlation between recovered sMRI components and ground-truth ones across settings for pICA and spICA were 0.73 and 0.96 (Fig. 5(d)), respectively. Recovered sMRI components had Hoyer indices of 0.36 and 0.79 for pICA and spICA (Fig. 5(d)), respectively.

**Fig. 5.**
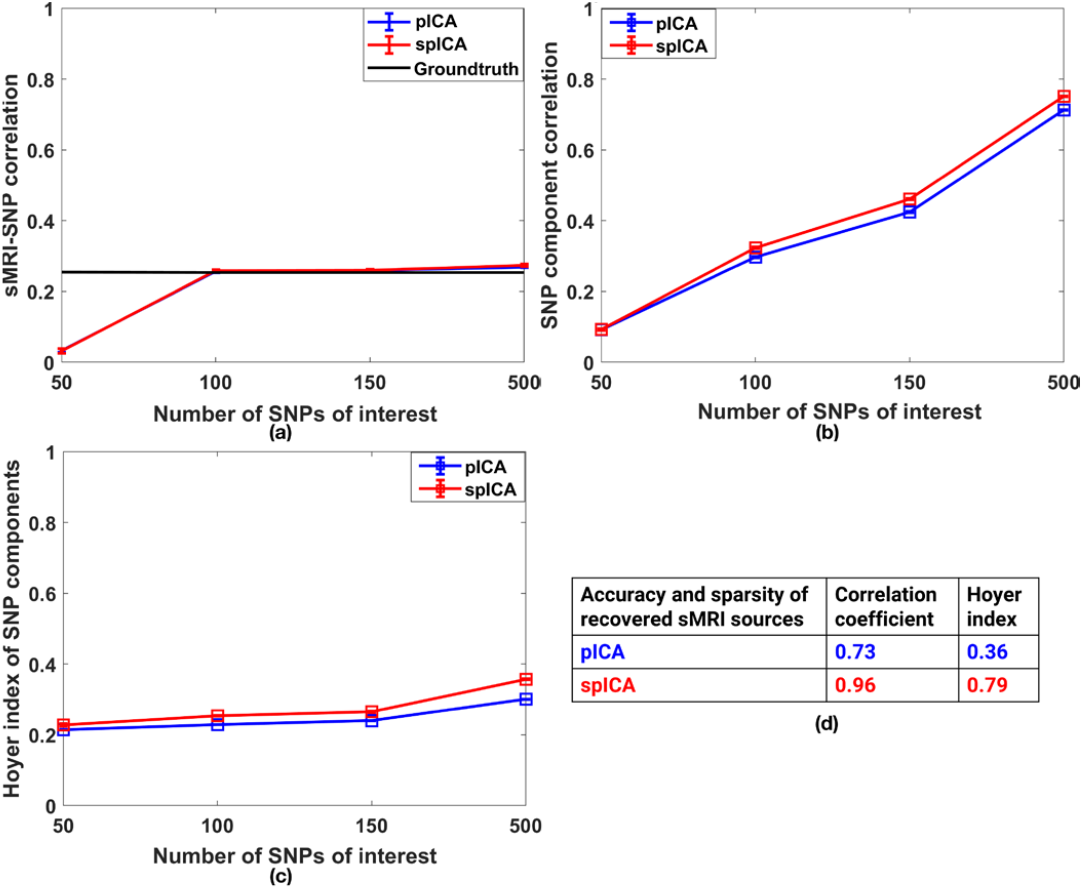
Scenario 3: Varying number of SNPs of interest from 50 to 500, the recovered (a) sMRI-SNP correlations (the ground-truth was 0.25 (black)), (b) SNP component accuracy, (c) Hoyer value of SNP components by spICA (red) and pICA (blue) and (d) sMRI component accuracy and sparsity. Compared to pICA, spICA recovered sMRI components with much higher accuracy and sparsity, and consistently higher SNP component accuracy and sparsity. The desired ratio between the number of SNPs of interest and total SNP number should be larger than 50/10000 for both pICA and spICA to recover the designed SNP components.

Fig. 6 and Fig. 7 present results under scenario 4 with the ground-truth correlation of 0.25. Fig. 6 displays the recovered sMRI-SNP correlations. As expected, performances from both spICA and pICA indicated that the larger the SNP pattern strength and the larger the PSNR value, the more accurate the detected sMRI-SNP association. The sMRI-SNP association accuracy is fairly stable with respect to PSNR, dropping with PSNR < 9dB. Under most conditions (i.e., SNP pattern strengths = 4, 5), the sMRI-SNP correlations identified by spICA were much closer to the ground-truth value of 0.25, as compared to those from pICA. When SNP pattern strength was 2, both spICA and pICA failed to detect the designed association.

**Fig. 6.**
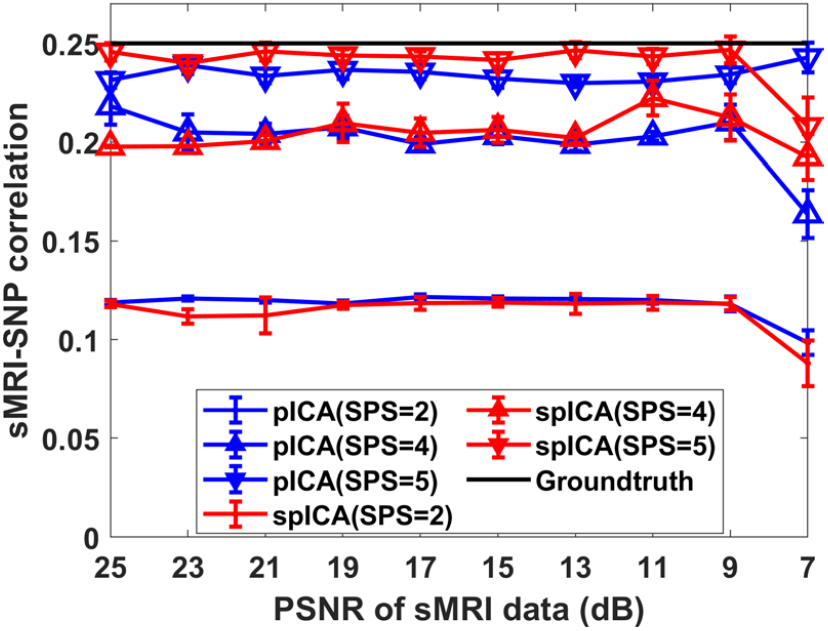
Scenario 4: The sMRI-SNP correlations detected by spICA (red) and pICA (blue) while varying noise levels of sMRI components and SNP pattern strength (SPS). The ground-truth of the correlation was 0.25 (black). The markers dot, upward triangle, and downward triangle represent results for SNP data with pattern strength of 2, 4, and 5, respectively. Importantly, compared to pICA, spICA yields higher accuracy in recovering the designed sMRI-SNP correlations when PSNR of sMRI components is greater than 7dB and when SNP pattern strength is 4 and 5.

**Fig. 7.**
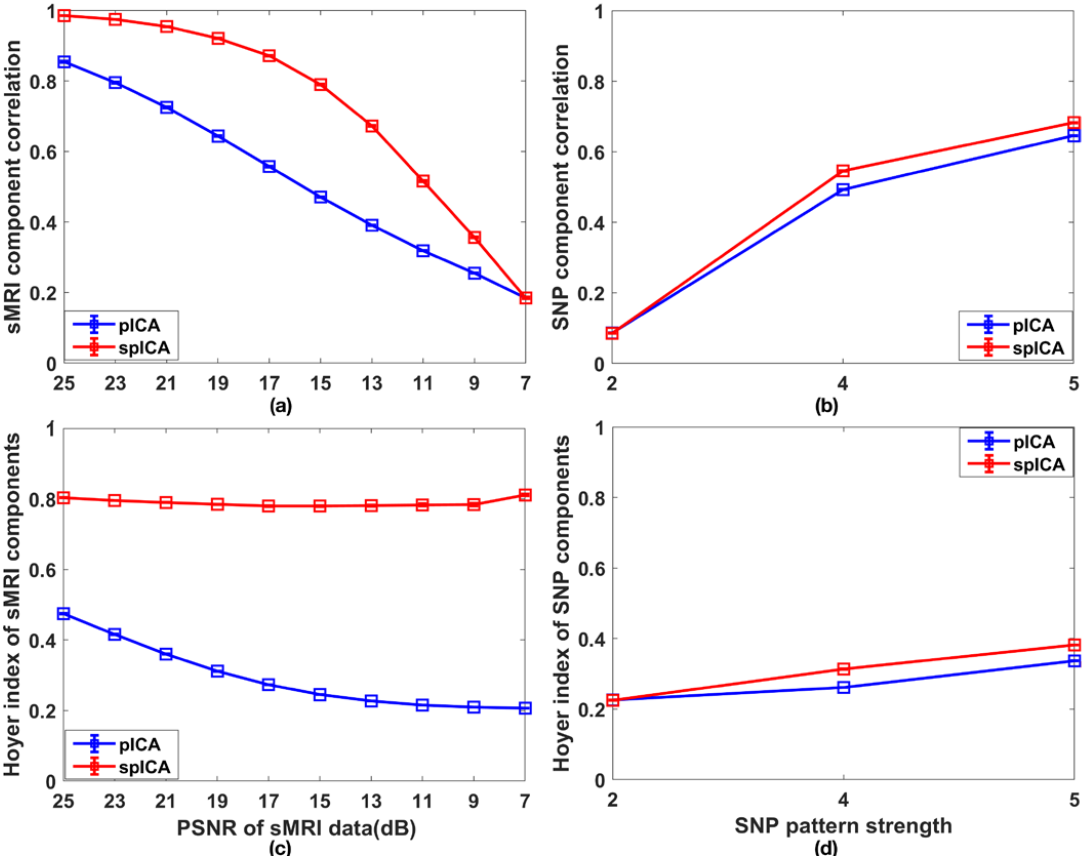
Performance of spICA and pICA while varying PSNR values of white Gaussian noise superimposed on sMRI components and SNP pattern strength: (a) accuracy and (c) sparsity of recovered sMRI components; (b) accuracy and (d) sparsity of recovered SNP components (the designed sMRI-SNP association was fixed at 0.25). Importantly, compared to pICA, spICA recovered sMRI components with much higher accuracy and sparsity, and consistently higher SNP component accuracy and sparsity. The desired SNP pattern strength should be larger than 2 for pICA and spICA to recover SNP components.

The accuracy of sMRI and SNP components (correlations with ground-truth components), and the sparsity of sMRI and SNP components (Hoyer indices) are displayed in Fig. 7. Fig. 7 (a) and (c) plot the detection accuracy and Hoyer indices of sMRI components across PSNR settings. Fig. 7 (b) and (d) plot the detection accuracy and Hoyer indices of SNP components across different SNP strength settings. Varying the strength of SNP patterns has a negligible influence on sMRI component recovery. Similarly, varying PSNR values of sMRI components has a negligible influence on SNP component recovery. Fig. 7 (a) and (c) demonstrated that as PSNR decreased, the accuracy of sMRI components decreased for both spICA and pICA, while the sparsity of sMRI components decreased for pICA but not so much for spICA. Nonetheless, spICA always yielded higher recovery accuracy and sparsity compared to pICA when PSNR was larger than 7. Fig. 7 (b) and (d) indicate that as the strength of SNP pattern increased, accuracy and sparsity of SNP components increased for both spICA and pICA. In addition, spICA outperformed pICA in terms of both accuracy and sparsity when the SNP pattern strength was larger than 2.

## IV. Results of spica on ukb smri and snp data

### 1) spICA identified three stable and replicable GMV-SNP component pairs

Using spICA, we identified that three GMV components (GMV IC1-3) consistently and significantly associated with one SNP component in all five spICA results (Fig. 8, partition 1-5). The spatial maps (brain regions/SNP loci) of highlighted GMV and SNP components were highly similar in five spICA results: correlation coefficients for GMV components = 1, correlation coefficients for SNP component > 0.77, and all the identified SNP components highlight SNPs in chromosome 17 (see Fig. 9 right panel and Supplemental Fig. S1). For simplicity, partition 1 was selected as the base (“final”) partition for assessing the stability of the identified GMV-SNP pairs. All the identified three GMV-SNP association pairs were significantly replicated in all five corresponding replication datasets.

**Fig. 8.**
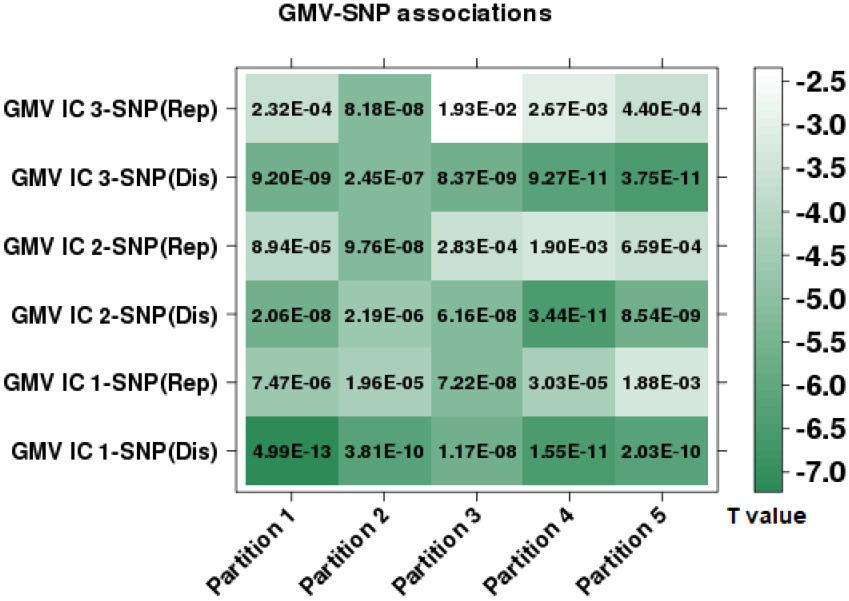
Stability and replicability of identified GMV-SNP pairs reflected by associations between identified three GMV components (IC 1-3) and one SNP component in discovery and replication datasets from 5 partitions (partition 1-5). The sea-green color is coded for association t values, and darker colors represent greater absolute t values (i.e., stronger associations). Association p values (in scientific format) were listed as text in each grid. ‘Dis’ and ‘Rep’ denote discovery and replication datasets, respectively (the same for Fig. 9 and Fig. 10).

**Fig. 9.**
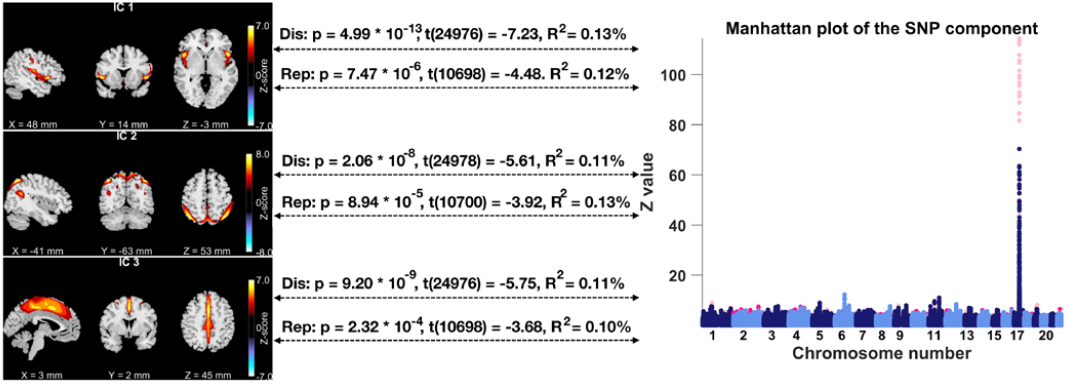
Three GMV components (left panel, |*z*| > 2.5) were significantly and negatively associated with one SNP component (right panel, the Manhattan plot). These associations were replicated in the replication dataset. In the Manhattan plot, hot colors represent positive weights, and cold colors denote negative weights, and the spike in chromosome 17 contains top SNPs related to the three GMV components.

Fig. 9 displays GMV-SNP association results of partition 1, where three GMV components (IC 1-3) were significantly and negatively associated with one SNP component highlighting SNPs in chromosome 17 (GMV IC 1-SNP *t* (24976) = -7.23, GMV IC 2-SNP *t* (24978) = -5.61, GMV IC 3-SNP *t* (24976) = -5.75), and these associations were replicated in the replication dataset (GMV IC 1-SNP *t* (10698) = -4.48, GMV IC 2-SNP *t* (10700) = -3.92, GMV IC 3-SNP *t* (10698) = -3.68). Highlighted brain regions were the superior temporal gyrus (GMV IC 1), superior/inferior parietal lobule and precuneus (GMV IC 2), cingulate and medial frontal gyri, and paracentral lobule (GMV IC 3).

### 2) Cognitive and neuroticism relevance of spICA-identified GMV and SNP components

The identified GMV components were significantly associated with multiple cognitive measures in discovery samples after FDR at p < 0.05 corrections for 20 tests (Fig. 10 (a)). Specifically, higher GMV in the superior temporal gyrus (GMV IC 1) was related to higher fluid intelligence score, greater maximum digits remembered correctly (numeric memory test), and shorter mean time to correctly identify matches (reaction time test), and these associations were replicated in the replication dataset. Furthermore, the associations between GMV in the superior temporal gyrus (GMV IC 1) and fluid intelligence score and maximum digits remembered correctly were stronger in older participants compared to younger ones in both discovery and replication datasets (Fig. 10 (b)). The association between GMV in the superior temporal gyrus (GMV IC 1) and mean time to correctly identify matches was stronger in younger individuals compared to older ones in the discovery dataset (Fig. 10 (b)). Moreover, higher GMV in the superior temporal gyrus (GMV IC 1) was related to smaller neuroticism score in the discovery dataset. Higher GMV in superior/inferior parietal lobule and precuneus (GMV IC 2) was related to lower fluid intelligence scores in the discovery dataset. Higher GMV in cingulate, medial frontal gyri and paracentral lobule (GMV IC 3) was associated with shorter mean time to correctly identify matches in both discovery and replication datasets. The identified SNP component was not significantly related to any cognitive measures tested.

**Fig. 10.**
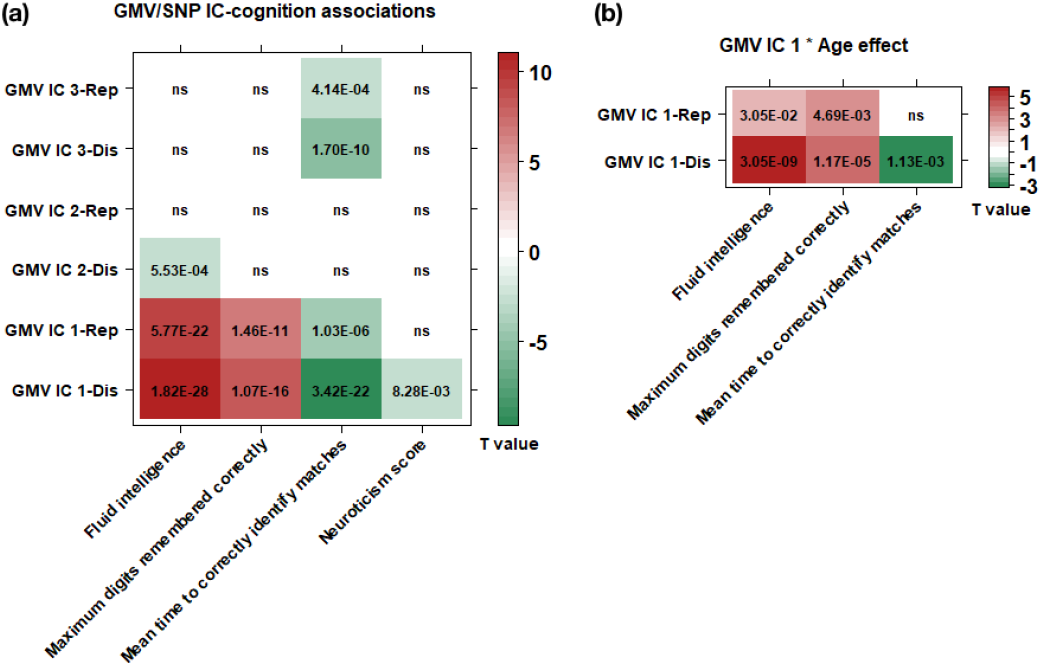
(a) Associations between identified GMV components and cognitive performance. (b) GMV IC 1 × age interaction effect for cognition prediction. The firebrick and sea-green colors are coded for positive and negative association t values, respectively, where darker colors represent greater absolute t values (stronger associations). Association p values (in scientific format) were listed as text in each grid.

### 3) Biological relevance of spICA-identified SNP component

The identified SNP component highlighted 790 SNPs in chromosome 17 (|z|>3.5, supplemental excel file). These 790 SNPs were mapped to 128 unique genes/long non-coding RNAs (lncRNAs), which were enriched in several canonical pathways such as oncostatin M signaling (p = 6.72×10^−5^), PPAR signaling (p = 2.01×10^−4^), IL-22 signaling (p = 2.40×10^−4^), role of JAK family kinases in IL-6-type cytokine signaling (p = 2.71×10^−4^), and thrombopoietin signaling (p = 3.00×10^−4^). The highlighted genes were enriched in inflammatory disease (p value range: [5.57×10^−5^, 1.01×10^−2^], 12 molecules) and inflammatory response (p value range: [5.57×10^−5^, 1.01×10^−2^], 15 molecules) pathways. The mapped 128 genes were not enriched in astrocytes/endothelial/microglia/neurons/oligodendrocytes cell-type specific gene sets identified in Zhang’s study [31].

Based on expression (eQTL), transcript expression (tQTL), and isoform percentage (isoQTL) quantitative trait loci summary statistics for the prefrontal cortex available on the PsychENCODE website (http://resource.psychencode.org/), we identified that out of 790 top SNPs, 207 acted as cis-eQTLs of 34 unique protein-coding genes and 17 lncRNAs, 175 were cis-isoQTLs of 78 unique transcripts, and 170 were cis-tQTLs of 69 unique transcripts. Based on methylation QTL (mQTL) summary statistics in the human frontal cortex provided in Jaffe’s study[30], we identified that 188 out of 790 top SNPs significantly regulated methylation levels of 141 unique CpG sites (see eQTL, tQTL, isoQTL and mQTL results in the supplemental excel file). Moreover, the regulation effects through eQTL/isoQTL/tQTL/mQTL were significantly enriched (*p* < 1.00×10^−4^).

## V. Discussion and conclusion

In this study, we propose a multivariate fusion method, spICA, for imaging-genetic studies. While building upon pICA for simultaneous independence maximization (via infomax ICA) and correlation optimization, spICA seeks further improvement in performance by leveraging the sparsity nature of sources. The sparsity constraint is implemented through a nonlinear Hoyer projection, which is applied directly to the estimated sources to suppress noise (background) and enhance the signal. Additionally, the sparsity enhancements are continually propagated by passing the reconstructed, cleaner data to the next iteration, allowing to tune the sparsity of sources effectively and explicitly.

Simulation results demonstrate that spICA outperformed pICA with improved accuracy for sMRI-SNP association detection and component spatial map recovery, as well as with enhanced sparsity for sMRI and SNP components under noisy case (scenario 4, heavy noise (small PSNR values) on sMRI data and small SNP pattern strength). Under scenarios 1-3 that a range of sMRI-SNP correlation strength, sample size, SNP source sparsity are examined, both spICA and pICA largely recover the designed sMRI-SNP linkage, but spICA outperforms pICA in terms of component spatial map recovery and sparsity of sMRI and SNP components.

Applying spICA to whole-brain GMV and whole-genome SNP data of 24985 unrelated European Caucasians recruited in UKB, we identify three GMV-SNP pairs passing Bonferroni correction, which are stably identified from all five discovery datasets and replicated in all five replication datasets. The identified GMV components highlight superior temporal, superior/inferior parietal, and medial frontal regions. The identified SNP component emphasizes SNPs in chromosome 17. The highlighted GMV components are significantly related to multiple cognitive measures such as fluid intelligence, maximum digits remembered correctly (numeric memory test), and mean time to correctly identify matches (reaction time test). More importantly, associations between superior temporal GMV and fluid intelligence and maximum digits remembered correctly increase with age in both discovery and replication datasets, while the association between superior temporal GMV and mean time to correctly identify matches decreases with age in the discovery dataset. Highlighted SNPs are enriched in inflammatory disease and inflammatory response pathways, and their regulation effects through eQTL/isoQTL/tQTL/mQTL in the frontal region are significantly enriched.

## Data Availability

All data produced in the present study are available at https://www.ukbiobank.ac.uk/.

