## Supplement for "Sparse Parallel Independent Component Analysis and Its Application to Identify Stable and Replicable Imaging-genomic Association Patterns in UK Biobank": supplement_Duan_SPICA_UKB_sMRI_SNP_2022_IEEETMI.docx

**Supplemental Materials:**

Require: datasets $\mathbf{X}_{1},$ $\mathbf{X}_{2}\boldsymbol{,}$ maximum number of the iteration $T$

Initialize the regularizer$\lambda$; preset Hoyer thresholds $q_{1}$,$q_{2}$; Hoyer sparsity enhancement step size $\Delta_{h1}$,$\Delta_{h2}$; unmixing matrices $\mathbf{W}_{1}$,$\mathbf{W}_{2}$; modality stopping flags $f_{1}$= FALSE,$f_{2}$= FALSE; entropy drop flags introduced by correlation optimization: $f_{c1}$= FALSE,$f_{c2}$= FALSE; $t_{1}=1$,$t_{2}=1$.

1: While ($t_{1}<T$ & $f_{1}$= FALSE) or ($t_{2}<T$ & $f_{2}$= FALSE) do

2: If $t_{1}<T$ & $f_{1}$ = FALSE	--Infomax on modality 1

3:	Do an infomax update on$\mathbf{W}_{\boldsymbol{1}}$ using stochastic gradient descent

4:	If	$\left\| \mathbf{W}_{1, \mathbf{t}_{1}}\mathbf{-}\mathbf{W}_{1,\mathbf{t}_{1}\boldsymbol{-1}} \right\|_{2}^{2}\boldsymbol{<}\boldsymbol{10}^{\boldsymbol{-6}}$

5:	$f_{1}$ = TRUE

6:	If the slope of the entropy of this modality is smaller than -10^-5^

7:	$f_{c1}$ = TRUE

8: If $\mathbf{t}_{2}\boldsymbol{<}\mathbf{T}$ **&** $f_{2}$ = FALSE --Infomax on modality 2

9:	Do an infomax update on$\mathbf{W}_{\boldsymbol{2}}$ using stochastic gradient descent

10:	If	$\left\| \mathbf{W}_{2, \mathbf{t}_{2}}\mathbf{-}\mathbf{W}_{2,\mathbf{t}_{2}\boldsymbol{-1}} \right\|_{2}^{2}\boldsymbol{<}\boldsymbol{10}^{\boldsymbol{-6}}$

11:	$f_{2}$ = TRUE

12: If the slope of the entropy of this modality is smaller than -10^-5^

13:	$f_{c2}$ = TRUE

14: If $f_{c1}$ = TRUE or $f_{c2}$ = TRUE	--Reduce the regularizer $\lambda$

15:	$\lambda=0.9\times\lambda$

16: Compute the new source matrix $\mathbf{S}_{1}$ using the newly updated $\mathbf{W}_{1}$ as:$\mathbf{S}_{1}\mathbf{=}\mathbf{W}_{1}\mathbf{X}_{1}$

17: If $f_{1}$ = FALSE	--Sparsity optimization on modality 1

18:	For each source ($j$-th row) $\mathbf{S}_{1,j}$ in $\mathbf{S}_{1}$ :

9: 19:	If the Hoyer index of the source $\mathbf{S}_{1,j}$ is less than the preset Hoyer value $q_{1}$:

20:	Do Hoyer projection$\mathbf{S}_{H1,j}\mathbf{=}\mathrm{Hoyer}\left( \mathbf{S}_{1,j} \right)$ with Hoyer step size $\Delta_{h1}$

21:	Else:$\mathbf{S}_{H1,j}\mathbf{=}\mathbf{S}_{1,j}$

**spICA algorithm pseudocode**

22: If the slope of the entropy of this modality is smaller than -10^-5^

23:	$\Delta_{h1}$= ${0.98\times\Delta}_{h1}$	--Reduce the Hoyer step size $\Delta_{h1}$

24: Reconstruct the data $\mathbf{X}_{1}$using the cleaner source $\mathbf{S}_{H1}$: $\mathbf{X}_{1}= \mathbf{W}_{\boldsymbol{1}}^{\boldsymbol{+}}\mathbf{S}_{H1}$

25: Compute the new source matrix $\mathbf{S}_{2}$ using the newly updated $\mathbf{W}_{2}$ as:$\mathbf{S}_{2}\mathbf{=}\mathbf{W}_{\boldsymbol{2}}\mathbf{X}_{\boldsymbol{2}}$

26: If $f_{2}$ = FALSE	--Sparsity optimization on modality 2

27:	For each source ($j$-th row) $\mathbf{S}_{2,j}$ in $\mathbf{S}_{2}$

9: 28:	If the Hoyer index of the source $\mathbf{S}_{2,j}$ is less than the preset Hoyer value $q_{2}$:

29:	Do Hoyer projection$\mathbf{S}_{H2,j}\mathbf{=}\mathrm{Hoyer}\left( \mathbf{S}_{\boldsymbol{2},j} \right)$ with Hoyer step size $\Delta_{h2}$

30: Else:$\mathbf{S}_{H2,j}\mathbf{=}\mathbf{S}_{\boldsymbol{2},j}$

31: If the slope of the entropy of this modality is smaller than -10^-5^

32:	$\Delta_{h2}$= ${0.98\times\Delta}_{h2}$	--Reduce the Hoyer step size $\Delta_{h1}$

33:	Reconstruct the data $\mathbf{X}_{2}$using the cleaner source $\mathbf{S}_{H2}$: $\mathbf{X}_{2}= \mathbf{W}_{\boldsymbol{2}}^{\boldsymbol{+}}\mathbf{S}_{H2}$

34: If $f_{1}$ = FALSE or $f_{2}$ = FALSE --Correlation optimization on modalities 1 and 2

35:	Update $\mathbf{W}_{1}$ and/or $\mathbf{W}_{2}$ based on correlation optimization with the regularizer $\lambda$

36: Increase the step $t_{1}$by 1: $\mathbf{t}_{1}\boldsymbol{=}\mathbf{t}_{1}\boldsymbol{+1}$

37: ${Increase the step t_{2} by 1:\mathbf{t}}_{2}\boldsymbol{=}\mathbf{t}_{2}\boldsymbol{+1}$

**S1, Loadings of identified GMV and SNP components in the replication dataset:**

Given the spatial patterns (i.e., weight of each voxel/SNP) of identified GMV-SNP pairs, we compute the expression levels (i.e., loadings) of the identified GMV and SNP components across individuals in the replication dataset using a projection method [1]: let $\mathbf{S}_{\mathbf{di}}$ and $\mathbf{S}_{\mathbf{dg}}$ denote the source/component matrices decomposed from the *discovery* GMV and SNP data, respectively. Let $\mathbf{X}_{\mathbf{ri}}$ and $\mathbf{X}_{\mathbf{rg}}$ represent *replication* GMV and SNP data, respectively. Since the GMV data was *not* optimized for sparsity, then the corresponding loading matrix of replication GMV data can be estimated as $\mathbf{A}_{\mathbf{ri}}=\mathbf{X}_{\mathbf{ri}}\boldsymbol{\times}\mathbf{S}_{\mathbf{di}}^{\mathbf{+}}$. On the other hand, recall that spICA utilized nonlinear sparsity regularization and reconstructed SNP data with cleaner sources at each iteration (i.e., denoising the SNP components of the discovery dataset). Thus, in this case, we reconstructed the loading matrix of SNP data using Tikhonov-regularized least squares [2] to account for the denoising effect in the replication SNP data. The loading matrix of SNP data was reconstructed as $\mathbf{A}_{\mathbf{rg}}^{\top}\mathbf{=}{\mathbf{(S}_{\mathbf{dg}}\mathbf{S}_{\mathbf{dg}}^{\top}\text{+}\text{α}\text{I}\mathbf{)}}^{\boldsymbol{-1}}\boldsymbol{\times}\mathbf{S}_{\mathbf{dg}}\mathbf{X}_{\mathbf{rg}}^{\top}$, where $\text{I}$ is an identity matrix, $\text{α}$ is a parameter calculated as in [3] to balance between retaining the signal and reducing the noise.

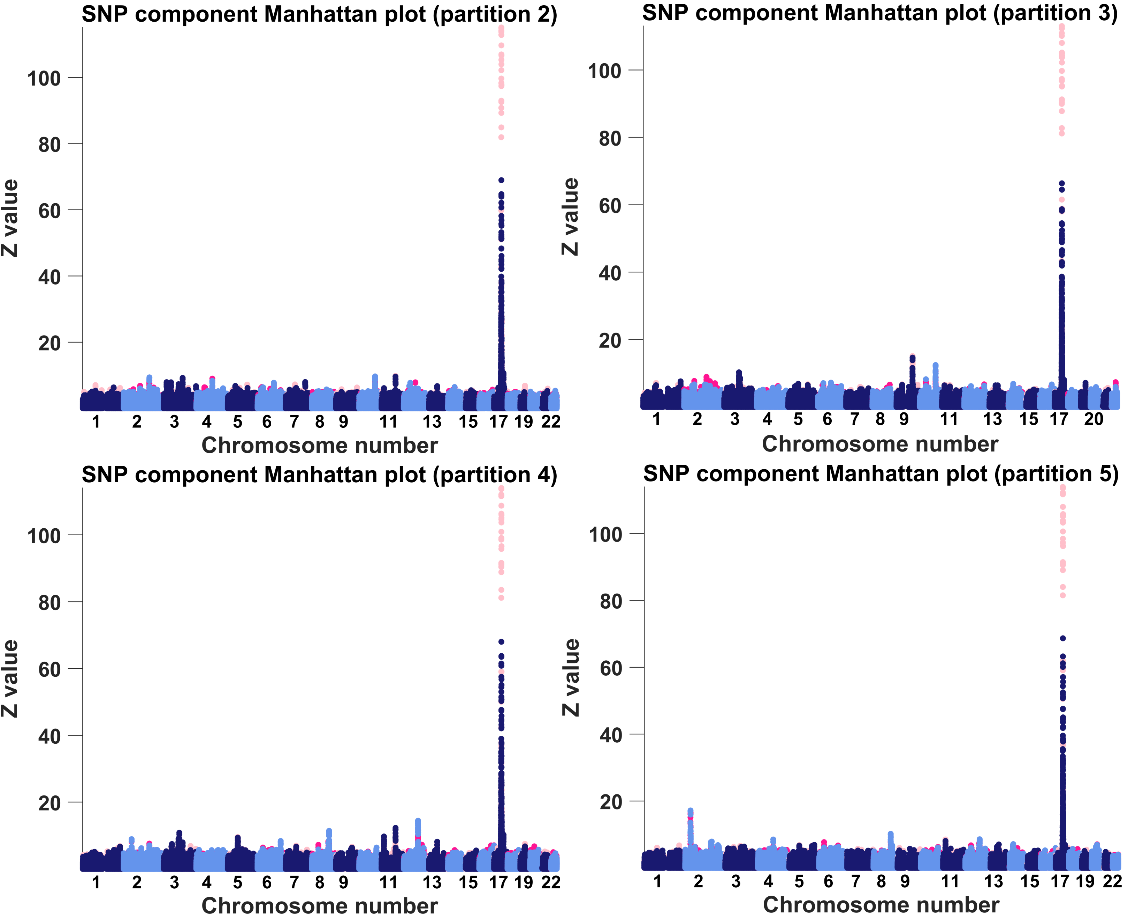

Fig. S1, Manhattan plots of spICA-identified SNP components from partitions 2-5. Note, hot colors represent positive weights, and cold colors denote negative weights.

TABLE S1. Summary of cognitive measurements and neuroticism score.

| Variables | Discovery (#) | Replication (#) | | Discovery (mean±std) | | | Replication (mean±std) |
| --- | --- | --- | --- | --- | --- | --- | --- |
| Fluid intelligence | 23071 | | 9887 | | 6.68± 2.05 | 6.67± 2.05 | |
| Maximum digits remembered correctly | 16838 | | 7257 | | 6.71± 1.48 | 6.67± 1.53 | |
| Number of incorrect matches in round | 23483 | | 10071 | | 3.58± 2.87 | 3.61± 2.86 | |
| Mean time to correctly identify matches | 23373 | | 10017 | | 593.61±109.04 | 591.73± 106.37 | |
| Neuroticism score | 21106 | | 9033 | | 3.80±3.16 | 3.92± 3.22 | |

Note, std denotes standard deviation.

[1]	J. Chen *et al.*, "G-protein genomic association with normal variation in gray matter density," *Hum Brain Mapp,* vol. 36, no. 11, pp. 4272-86, Nov 2015, doi: 10.1002/hbm.22916.

[2]	G. Strang, *Computational science and engineering*. Wellesley-Cambridge Press, 2007.

[3]	K. K. Duan, R. F. Silva, J. Y. Chen, D. D. Lin, V. D. Calhoun, and J. Y. Liu, "Sparse Infomax Based on Hoyer Projection and Its Application to Simulated Structural Mri and Snp Data," (in English), *I S Biomed Imaging,* pp. 418-421, 2019. [Online]. Available: <Go to ISI>://WOS:000485040000091.
